# Assessment of fetal corpus callosum biometry by 3D super-resolution reconstructed T2-weighted MRI

**DOI:** 10.1101/2023.06.08.23291142

**Authors:** Samuel Lamon, Priscille de Dumast, Vincent Dunet, Léo Pomar, Yvan Vial, Mériam Koob, Meritxell Bach Cuadra

## Abstract

**Objective:** To assess the accuracy of corpus callosum (CC) and its sub-segments’ biometry by super-resolution (SR) 3-dimensional fetal brain MRI in comparison to measurements in 2-dimensional or 3-dimensional ultrasonography (US) and clinical low-resolution T2-weighted MRI sequences (T2WS).

**Method:** We performed fetal brain biometry of the overall length of the CC, the heights of its sub-segments and its area by two observers (one junior observer, obs1, and one senior pediatric neuroradiologist, obs2) in a cohort of 57 subjects (between 21 and 35 weeks of gestational age (GA), including 11 cases of partial agenesis of CC). Obs1 made measures on US, T2WS, and SR, and obs2 in T2WS and SR. Regression curves of CC biometry with GA were done. Statistical analysis of inter-modality (US vs. T2WS, US vs. SR, and T2WS vs SR) agreement for single observer (obs1) and inter-modality (US vs. T2WS, and US vs. SR) between observers (obs1 vs obs2) were also conducted.

**Results:** Our study shows a high concordance through GA of CC measurements performed by SR in comparison with US, with a higher agreement than biometry based on T2WS clinical acquisitions. For obs1, SR measurements are highly concordant to US (except for the genu and the CC length) and helps visualizing the splenium. For obs2, SR measurements are highly concordant to US, except for the rostrum and the CC length. Rostrum and Genu (forming the anterior callosum) are the subsegments with larger variability. Regression curves by SR overlay more accurately those from the literature (by US) for the CC length, the splenium and the body than T2WS.

**Conclusion:** Super-resolution MRI could be used in the biometrical assessment of the CC, providing measurements close to US, except for the anterior part of the CC Thanks to its 3D-visualisation capacity and improved through plane spatial resolution, it allows to perform CC biometry more frequently than on T2WS.

## 1 INTRODUCTION

The corpus callosum (CC) is the largest brain commissure connecting homologous structures of both cerebral hemispheres and is fully formed after complex embryogenesis steps in-utero at 20 weeks of gestation (1). The corpus callosum is divided into four parts: the rostrum, the genu, the body and the splenium, from the most anterior to the most posterior part. Complete or partial agenesis of the corpus callosum (cCCA and pCCA, respectively) are among the most frequent brain malformations prenatally detected, with an estimated prevalence of 0.3 to 0.7% in the general population and of 2 to 3% in people suffering from neurodevelopmental disorders (2). The neurodevelopmental outcome in children born with a CC anomaly, now grouped over the term failed commissuration, is extremely heterogeneous, ranging from normal neurodevelopment to severe delay, depending not only on the specific type of CC anomaly, but also on the presence or absence of potentially associated cerebral and/or extra-cerebral malformations (3). Today, the main challenge does not lie in establishing the diagnosis, but in estimating the neurodevelopmental prognosis which remains very difficult to predict, as shown by post-natal follow-up studies (4). Therefore, the assessment of the integrity of the fetal CC, with an accurate biometrical and morphological analysis, is crucial for the evaluation of the pre- and post-natal management and the prognosis (5).

Ultrasound (US) and Magnetic Resonance Imaging (MRI) are complementary methods for evaluating fetal brain structural development. At the occasion of the routine 2^nd^ trimester ultrasound examination, a screening for callosal abnormalities takes place. Direct evidence of a cCCA or pCCA, by absence of CC visualization, completely or partially, respectively, usually requires a mid-sagittal image. As this plane is not included in the required planes for the screening, we usually rely on indirect signs of CC absence on an axial image, such as the absence of the cavum of the septum pellucidum, that are more or less present according to the missing part (6). Anomalies of the CC also comprise the dysplasias, like the hypo- or hyperplasia where the CC is abnormally thin or thick, respectively. In case of a suspected callosal abnormality, an additional detailed US (a so-called neurosonogram, usually acquired transvaginally), is eventually performed by an expert and relies on the 3D imaging and sagittal, axial, and coronal views. Structural magnetic resonance T2-weighted sequences (T2WS) are recommended at 32 weeks of GA as a complement to the targeted US to either confirm and characterize or to rule out a suspected callosal abnormality and to look for other cerebral malformations (7,8). These two imaging modalities have their strengths and weaknesses and the study of the CC remains their biggest source of disagreement (3). Even if debated (9), mainly because of the lack of clarity regarding the way US exams were performed, a recent large prospective multicenter study, the MERIDIAN study, showed that, in failed commissuration, MRI was more accurate than US in detecting and characterizing CC anomalies, and this changed the prognosis and the clinical management in around 45% of the cases (3,10). Indeed, while US has better spatial resolution than MRI and benefits from 3D reconstruction techniques (11), it may be limited by the position of the fetus, mother habitus or oligohydramnios for example. However, without contesting the benefit of MRI, some authors balanced the sonographer expertise over the contribution of MRI (6). Alike 3D- US, MRI allows a multiplanar acquisition. Its main strength is a good tissue contrast. However, clinical T2WS MRI acquisitions are still highly sensitive to motion and are limited by the acquisitions in thick slices (around 3 mm through plane) as major drawback. Consequently, while a sagittal plane depicting the CC, the aqueduct of Sylvius and the pituitary gland is mandatory on fetal brain MRI, images are not always in the right anatomical plane, which may lead to important approximation errors in the fetal CC shape analysis and measurements (12). Postnatally, MRI remains the gold standard for pathological CC analysis, often supporting the reclassification of cCCA into pCCA (7) or showing additional brain abnormalities that may change the prognosis (1,6).

Super-resolution (SR) reconstruction of fetal brain MRI aims at overcoming the limitations of clinical low-resolution fetal MRI series, mainly motion artefacts and low spatial resolution. SR is a post-processing technique based on inter-slice motion correction and scattered data interpolation methods (13–19), where, by incorporating multiple orthogonal scans with thick slices, a volumetric motion free high-resolution image can be estimated. When SR succeeds, this allows to view perfect orthogonal planes in a 3D MR volume, with an isotropic resolution between 0.5 and 1.2 mm (3). The multiplanar reconstruction (MPR) in any plane is of particular interest in the accurate measurement of the CC and its potential abnormalities. However, like any other reconstruction method, SR may distort the real anatomy (3). Thus, previous works have explored the value of SR reconstruction to perform fetal brain biometry, for whole brain and posterior fossa (20–22) in comparison to clinical T2WS MRI, ocular biometry (23), as well as in normative fetal brain atlases (12,24). To our knowledge, apart from the length of the CC, detailed CC biometry has not yet been evaluated on SR reconstructed fetal brain MRI. Therefore, SR must be validated in comparison to the reference standard for fetal CC biometry used at the moment, i.e. US, before its use and dissemination in clinical practice. The primary purpose of this study is to assess whether SR is closer to US than T2WS in the measurement of the normal corpus callosum and its sub-segments. A secondary objective aims at exploring whether SR can assess pCCA.

## 2 MATERIALS AND METHODS

### 2.1 Cohort

We retrospectively utilized fetal brain MRI exams performed from 2014 to 2021 at our institution, the Lausanne University Hospital. All of them were conducted on medical indication. From this database, we selected 62 MRI exams with at least 3 orthogonal T2WS series: 50 were considered normal (either no anomaly was detected, or they presented a mild ventriculomegaly, i.e., <12mm) and 12 presented pCCA (defined by absence in whole or in part of one or more sub-segments of CC and is recognized by a short or abnormal shape of CC).

In the normal patients’ cohort, three exams were discarded because of a twin pregnancy (creating a too important risk of confusion between the fetuses) and one due to an opposition to the general consent form. In the pCCA patients’ cohort, one exam was discarded because of a twin pregnancy (see Figure 1). Consequently, a total of 57 T2WS exams were included in this study, where 46 were considered normal (38 without anomaly and 8 with mild ventriculomegaly) and 11 presented pCCA. Gestational age (GA) ranged from 21 to 35 weeks in the normal group and from 22 to 31 weeks in the pCCA group. GA determination was based on the last menstrual period and confirmed by at least one US examination during the first trimester of pregnancy. All images were anonymized prior to further analysis. This retrospective study was part of a larger research protocol at our institution approved by the local ethics committee (CER-VD 2021-00124).

**Figure 1.**
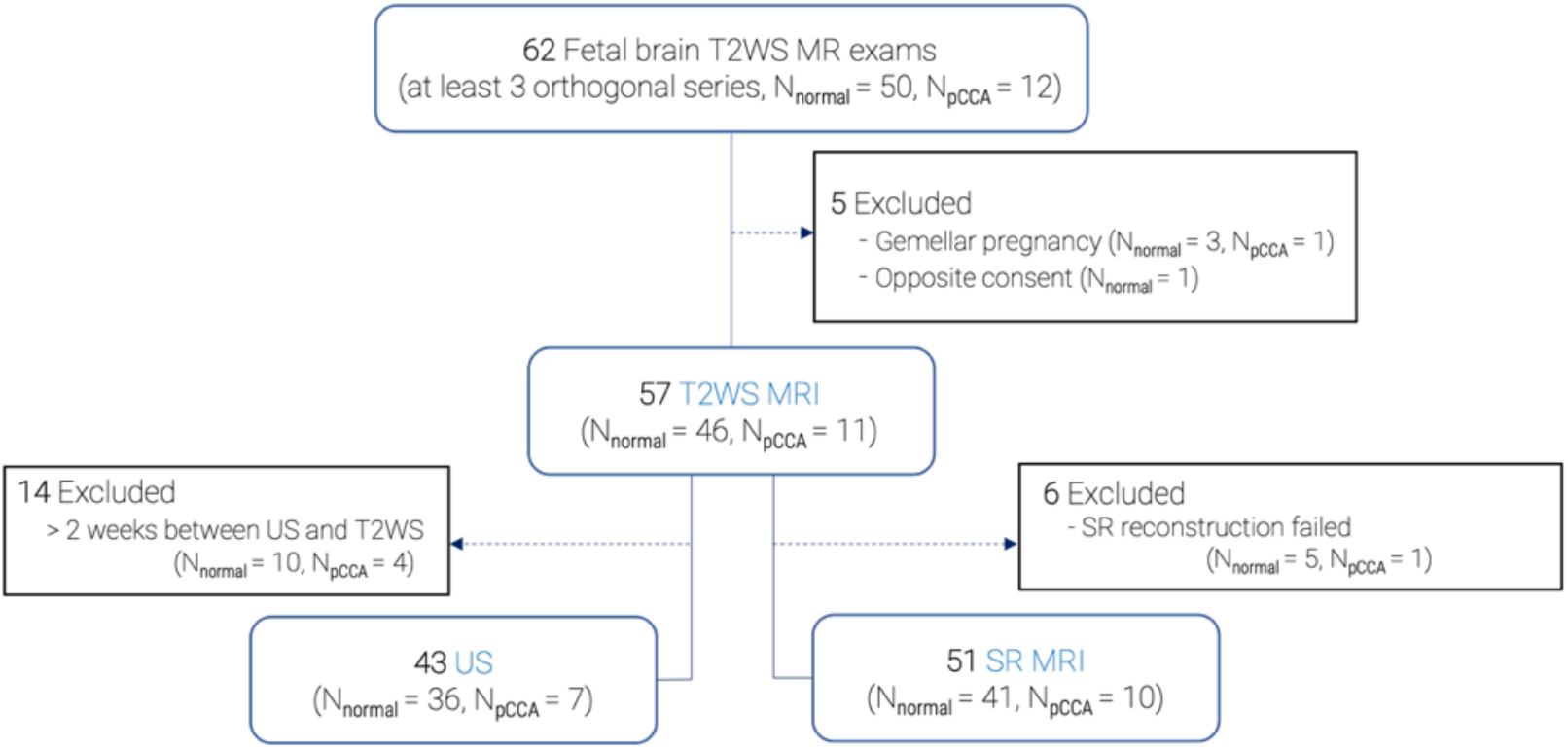
Data selection flow chart.

### 2.2 Magnetic Resonance Imaging

#### 2.2.1 Clinical acquisitions

Clinical MR images were acquired either at 1.5 T (MAGNETOM Aera, Siemens Healthcare, Erlangen, Germany) (91% of the exams) or at 3 T (MAGNETOM Skyrafit, Siemens Healthcare, Erlangen, Germany) (9% of the exams). The fetal brain MRI protocol included T2-weigthed Half-Fourier Acquisition Single-shot Turbo spin Echo (HASTE) sequence in the three orthogonal orientations. On average, for each subject, 7 T2WS series (also denoted as stacks) were available (range of 3 to 20) and the best stack was visually selected according to its quality. Figure 2a (middle row) shows an example of one T2WS HASTE sagittal acquisition (good in-plane sagittal resolution but thick slices visible in coronal and axial views), where the sagittal view has been rotated for better visualization.

**Figure 2.**
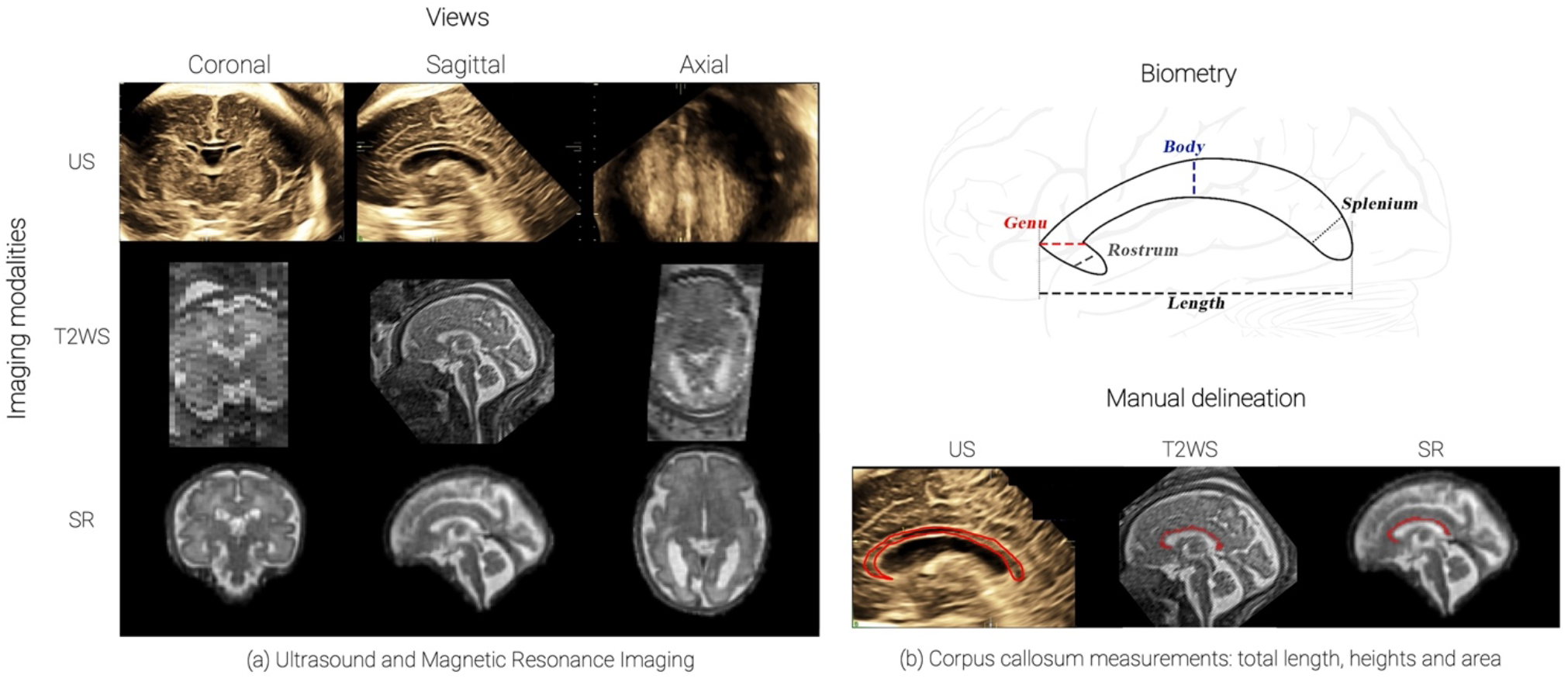
Imaging of the Corpus Callosum (CC): (a) Examples of the different imaging methods: a 3D ultrasound (top row), one T2WS MRI with thick slice series acquired in sagittal orientation (middle row, sagittal view has been manually reoriented for better visualization, additional low-resolution T2WS in other orientations, coronal and axial, are also acquired), 3D super-resolution (SR) reconstruction MRI (bottom row); (b) Biometry of the CC and its sub-segments (from anterior to posterior : rostrum, genu, body, and splenium) and manual delineation of CC area (bottom).

#### 2.2.2 Super-resolution reconstruction

The super-resolution reconstruction pipeline consists, in order, in the selection of low-resolution T2WS stacks, brain extraction, bias field correction, inter-slice motion estimation based on slice-to-volume rigid registration, and SR reconstruction. We used the MIALSRTK pipeline (14) which is based on an inverse problem formulation that is solved via an efficient total variation regularization. The selection of the series used for the SR reconstruction was done based on visual inspection, and T2WS stacks that exhibited extreme level of motion and/or intensity signal dropouts (thus, that were not exploitable for radiological reading neither) were excluded. The automated brain extraction was manually corrected if needed. In our study, on average 5.6 T2WS stacks per subject’s reconstruction were used (range of 3 to 9). All SR images were reconstructed with an *isotropic spatial resolution* matching its input in-plane resolution (in average of around 1.1 *mm*^3^ for 1.5T and 0.5 *mm*^3^ for 3T exams). Figure 2a (bottom row) shows an example of SR image. Six MR exams failed to be reconstructed due to either bad T2WS quality or strong motion remaining. A total of **51 SR exams** (41 normal and 10 pCCA) were finally available for measurements.

### 2.3 Ultrasound imaging

We selected US images within a time frame of two weeks before or after MR imaging, as recently suggested in the MERIDIAN cohort (25) and in many other studies (6,26,27). The precise time difference between imaging methods is illustrated in Supplementary Material (Figure S1). We believe that it is a good compromise to maintain a good framework for comparing biometric measurements while avoiding excluding too many patients. This led to 43 US sessions (36 normal and 7 pACC) available for performing measurements (Figure 2a, top row). The US images were acquired on General Electric (ZIPF, Austria) Voluson 730, E8, E10 devices, equipped with 5- to 8-MHz 3D transabdominal and transvaginal transducers. They have been acquired by members of the ultrasound and fetal medicine unit of our institution, either experimented obstetricians or midwives specialized in fetal neurosonography. Of all the US series used, 92.9% of them were 3D-US (39 out of 43) and only 3 out of 43 subjects (7.14%) were 2D-US. In the case of 3D-US, the acquisitions of the brain volumes containing the CC are performed after optimization of the 2D image starting from a bi-parietal diameter or a trans-cerebellar axial plane using a multiplanar mode with the best resolution available. The 3D volume is then displayed as multiple orthogonal 2D images, that are isotropic (28). In the case of 2D-US, they are acquired on a mid-sagittal plane starting from the same trans-cerebellar axial view, by aligning the transducer along the anterior fontanelle and the sagittal suture which serve as an acoustic window (29). Volume contrast imaging (VCI) is a tool that provides a thin 3D slice of the studied view. It helps reducing artifacts and increasing image resolution and contrast (28). In our study, this mode was used from 1 mm at 18 weeks up to 4 mm at 38 weeks.

### 2.4 Measurements

#### 2.4.1. CC biometry and manual delineation

Two observers, one junior observer (obs1) without specific experience in fetal brain MRI and one senior pediatric neuroradiologist (obs2), with 15 years of experience of fetal brain MRI, independently measured the overall length of the CC and heights of its sub-segments on both MRI datasets (T2WS and SR). Both were blinded for clinical data, including GA. Obs1 also performed the same measurements on US images, supervised by a midwife with 10 years of experience in fetal brain sonography and a skilled obstetrician with 35 years of experience, who both reviewed US measures at multiples time and validated them. Thus, US measurements will be used as *standard for reference* expert biometry.

On MR imaging (T2WS and SR), the free-resource ITK-SNAP software, version 3.6.0 (30) was used, and the images were then re-oriented to fit in the orthogonal axis. For T2WS, the best low-resolution stack for each orthogonal plane was chosen after a visual examination. On US, measurements were directly performed on the US devices, which allowed to re-navigate in all planes to find the best mid-sagittal one available. All CC measurements were related to the length and height of the hypoechoic area, excluding the boundary hyperechoic structures. They were performed with a 0.1 mm resolution cross-shaped caliper.

The following CC measurements (Figure 2b, top) were done on each imaging (US, T2WS, and SR): the outer-outer CC length (***LCC***), and the **heights of** the rostrum (***Rostrum***), genu (***Genu***), body (***Body***), and splenium (***Splenium***), according to the standard techniques described in the literature (31):

- *Rostrum* is the most anterior part which is oriented postero-inferiorly,
- *Genu* is defined as the segment situated anteriorly to a line passing through the anterior fornix and parallel to another line passing through the posterior fornix and the quadrigeminal plate,
- *Body* is situated between the splenium and the genu, and
- *Splenium* corresponds to the posterior 20% of the CC.

The measurements of pCCA followed the same methodology as for normal cases (examples of pCCA are illustrated in Figure 3).

**Figure 3.**
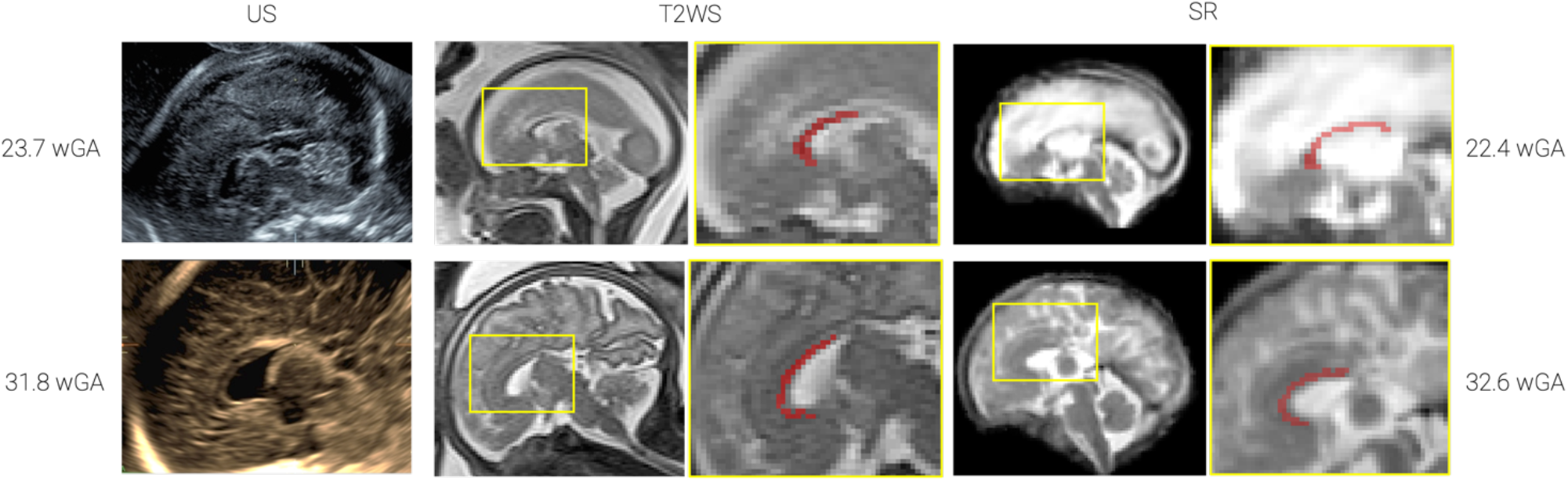
Examples of pCCA with the three imaging methods: patient (top row) is around 23 weeks of GA (23.7 wGA at time of US, and 22.4 wGA at time of MRI); patient (bottom row) is around 32 weeks of GA (31.8 wGA at time of US and 32.6 wGA at time of MRI).

The CC measurements were performed on the best midsagittal images available. First, all measurements were performed on T2WS as a block. Then, they were performed on US only, and afterwards on SR. For each imaging type, measurements lasted for approximately one week. CC measurements were always made three times in a row by each observer and were then averaged to minimize the intra-measurement variability. Remotely, we repeated the measurements on a subset of 10% of the exams (9 normal and 3 pCCA) serving as a reproducibility study.

Finally, manual delineation of the CC on T2WS and SR was done with the Paintbrush mode in ITK-SNAP open-source software (30). The CC area was approximated by the addition of all those voxels times its voxel size. On US, we manually drew the outer contour of the CC using the US device, which automatically gave a measure of the CC area (Figure 2b, bottom).

#### 2.4.2. Image Quality evaluation

For all modalities, image quality scores were estimated by obs1. T2WS quality criteria consisted of the mean of 6 items, three of them on the CC (quality of the visualization of the CC, visualization of a whole CC, amount of blurring of the CC), and three on the global stack quality (obliquity of the plane, global stack motion, global stack blurring). Each item could be rated from 0 to 3 (0=unusable, 1=bad, 2=average, 3=good). Similarly, for SR reconstruction, a quality assessment was done using the same criteria than the ones used for T2WS, though without the obliquity of the plane and the global stack motion, which are irrelevant with SR.

The quality of US images was scored using the quality score proposed by Pomar et al. (29) in their sonographic study of the CC. It consists of the sum of 5 criteria: strict sagittal plane with clear visibility of the cerebellar vermis, the brain stem, the fourth ventricle and the CC, the visualization of the 4 parts of the CC, a sufficient zoom, a clear differentiation of CC from the cavum septum pellucidum, and a right placement of the calipers. These criteria were rate as either 1=yes or 0=no. 5 points were equivalent to a good quality, 3 to 4 points to an average quality, 1 to 2 to a bad quality and 0 to an unusable exam.

### 2.5 Regression and statistical analysis

#### 2.5.1. Regression with gestational age

Regression analysis was done using measurements of normal subjects only (including mild ventriculomegaly and excluding pCCA). We compared our normative regression with previously validated published charts (31,32) serving as first assessment. Specifically, US measurements are compared to Pashaj et al. (31) for all CC biometric measurements. T2WS CC length is compared to Tilea et al. (32). To our knowledge, neither MR T2WS nor SR measures of CC sub-segments exist. CC area are compared to previously reported area from US imaging (33).

#### 2.5.2. Statistical analysis

We evaluate the discrepancies between the subsets of the measurements performed:

- We assess the discrepancies between the imaging setting (US, T2WS, and SR) measurements of obs1 (inter-modality agreement for a single observer).
- To remove the confounding of the experience, we assess the inter-modality agreement (US vs. T2WS, and US vs. SR), between observers, in which US measurements are those performed by obs1 that are validated by two US experts, and MR measurements (T2WS and SR) are from the MR experienced obs2.

For each of these comparisons, a paired Wilcoxon rank sum test was performed, with statistical significance set to p < 0.05. P-values are adjusted for multiple comparisons (five: ***LCC***, ***Rostrum***, ***Genu***, ***Body*** and ***Splenium***) using Bonferroni correction.

## 3 RESULTS

### 3.1 Analysis of different imaging datasets

Figure 4 summarizes the quality evaluation (range of blues) and the missing values (red) in the different evaluated imaging methods. Overall, missing values were more often occurring in US than in T2WS and SR. Out of 43 US exams, measurements were not possible in 14 cases (32.5%). In one half of them we observed a bad US image quality, in the other half an acceptable image quality and none of them occurred on an excellent quality exam. In T2WS, out of 57 exams, measurements were not possible 11 times (19.3%). Measurements were mostly missed for the rostrum’s height (10 cases out of 57, 17.5%), including 8/10 (80%) acceptable quality exams and 2/10 (20%) excellent quality exams. Finally in SR, only 2 cases out of 51 (3.92%) had missing measurements and they both involve the rostrum’s height (one on a bad quality exam and the other one on an acceptable quality exam). The higher overall quality of T2WS MRI over US is probably explained, at least partially, by the fact that subjects were initially selected on the basis of the MR exam.

**Figure 4.**
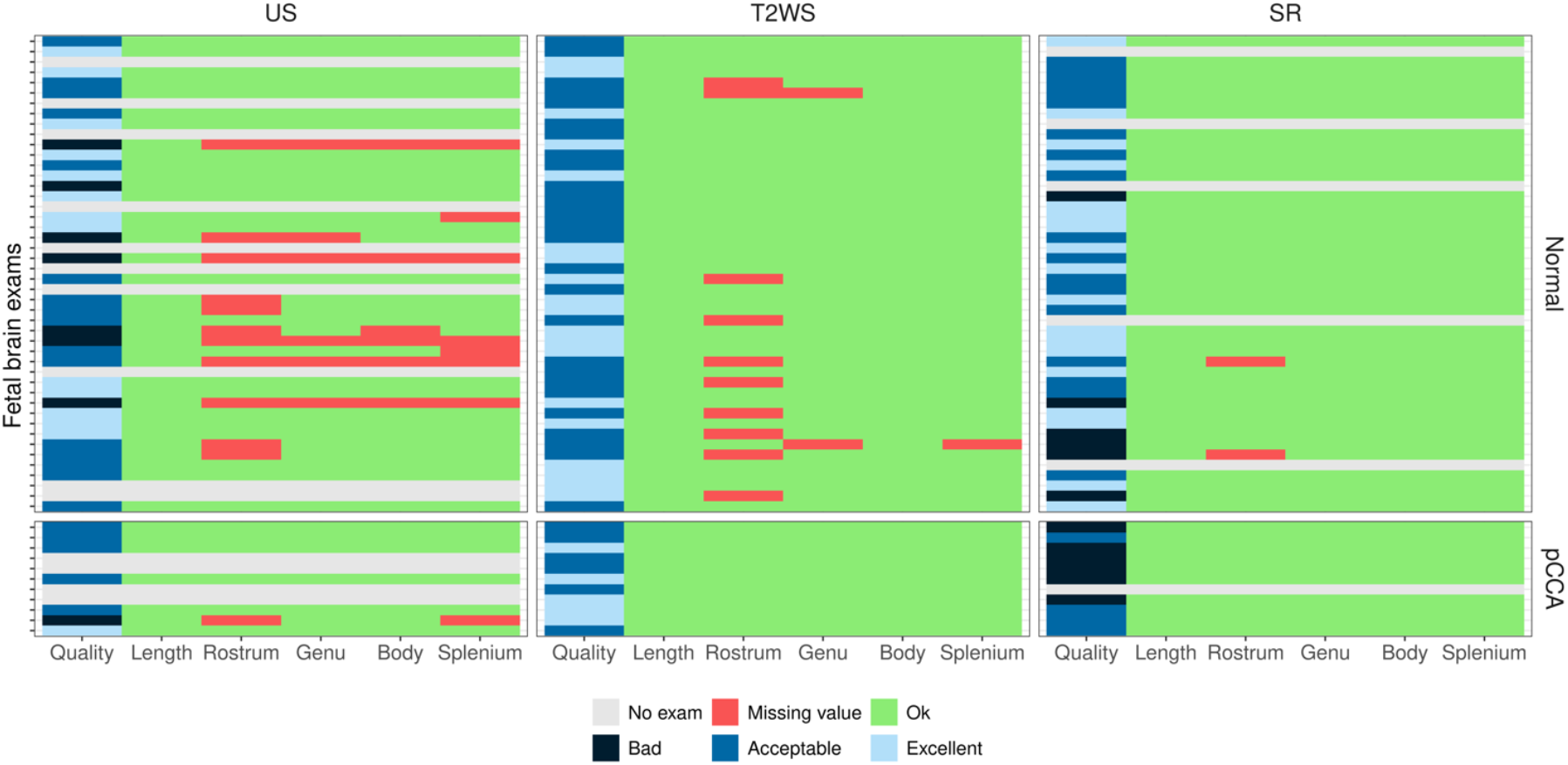
Imaging quality and availability of measurements.

### 3.2 Corpus callosum length and heights

Figure 5 shows the results of ***LCC*** measurements of intra-observer (obs1) on the three imaging techniques, US, T2WS and SR. Let us recall that pathological subjects with partial CCA are shown (illustrated by triangles) but not used for the regression curves. US and T2WS measurements are respectively compared with reported value in the literature from US (31) and T2WS MRI (32) (gray solid and dashed lines). In both cases, ***LCC*** shows a high agreement with previous works. The fourth panel in Figure 5 illustrates that the regression of the SR (yellow) overlayed on the regression of our measurements for US and T2WS. In fact, the SR measurements better fit the US (red) than T2WS (blue). The repeatability study on 12 subjects shows overall a high intra-class correlation coefficient of observer 1 (overall >0.99) in all three imaging settings (see Supplementary Material, Table S1).

**Figure 5.**
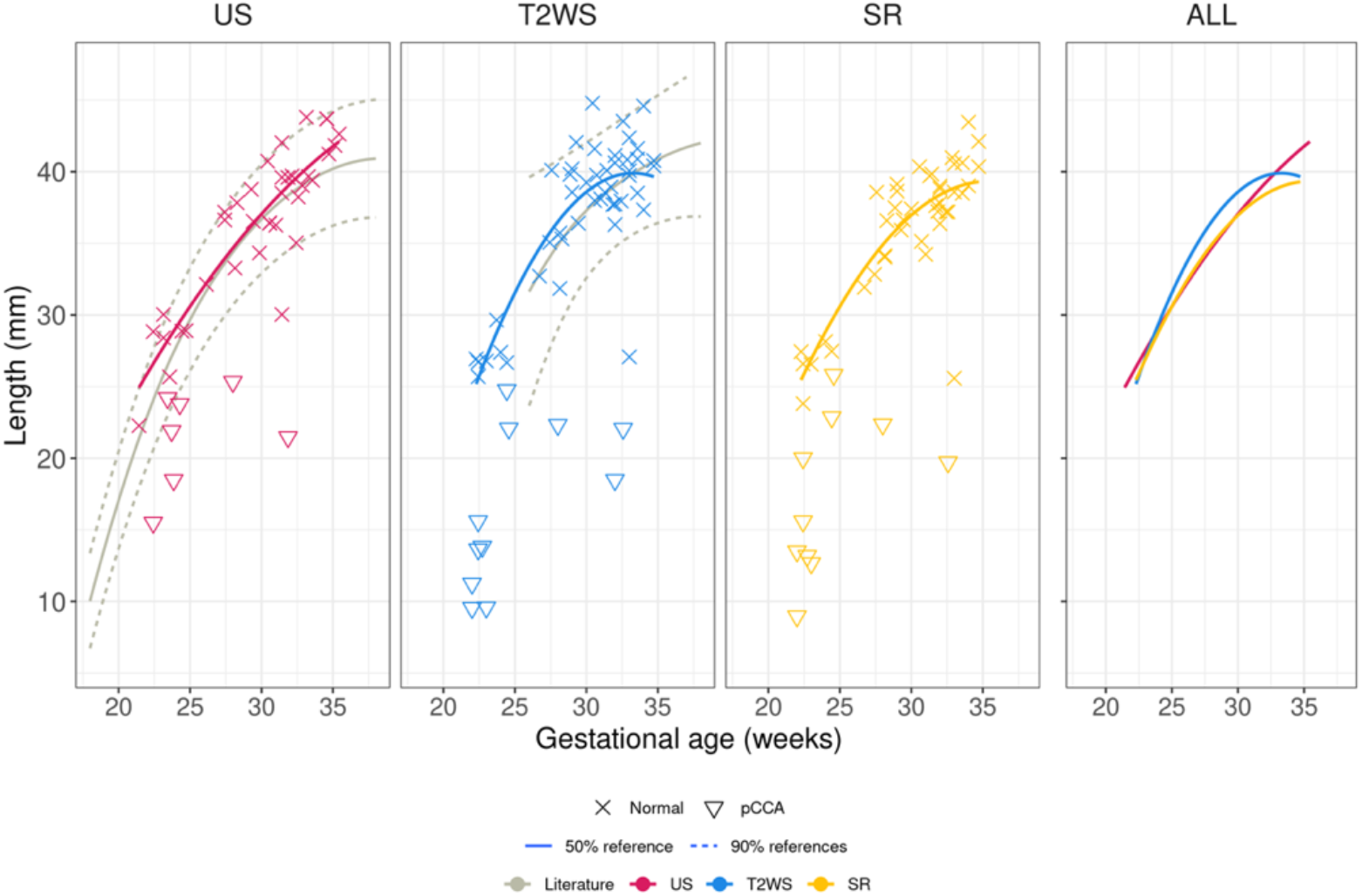
Length of the corpus callosum by obs1. US and T2WS measurements are respectively compared with previous reported value in the literature (gray solid and dashed lines) from US (31) and T2WS MRI (32). Pathological subjects with partial CCA are shown (illustrated by triangles) but not used for the regression curves.

**Figure 6** shows the comparison of CC sub-segments’ heights measurements done by obs1 on the three imaging setting US, T2WS and SR per each sub-segment. We also illustrate previous values reported in the literature based on the US only (31) as no reported sub-segment values based on MR T2WS nor SR exist to our knowledge. Overall, for all imaging US, T2WS and SR, the agreement with previous values reported in literature is high for ***Body*** and ***Splenium***, while it deviates more for ***Rostrum*** (US) and ***Genu*** (all imaging).

**Figure 6.**
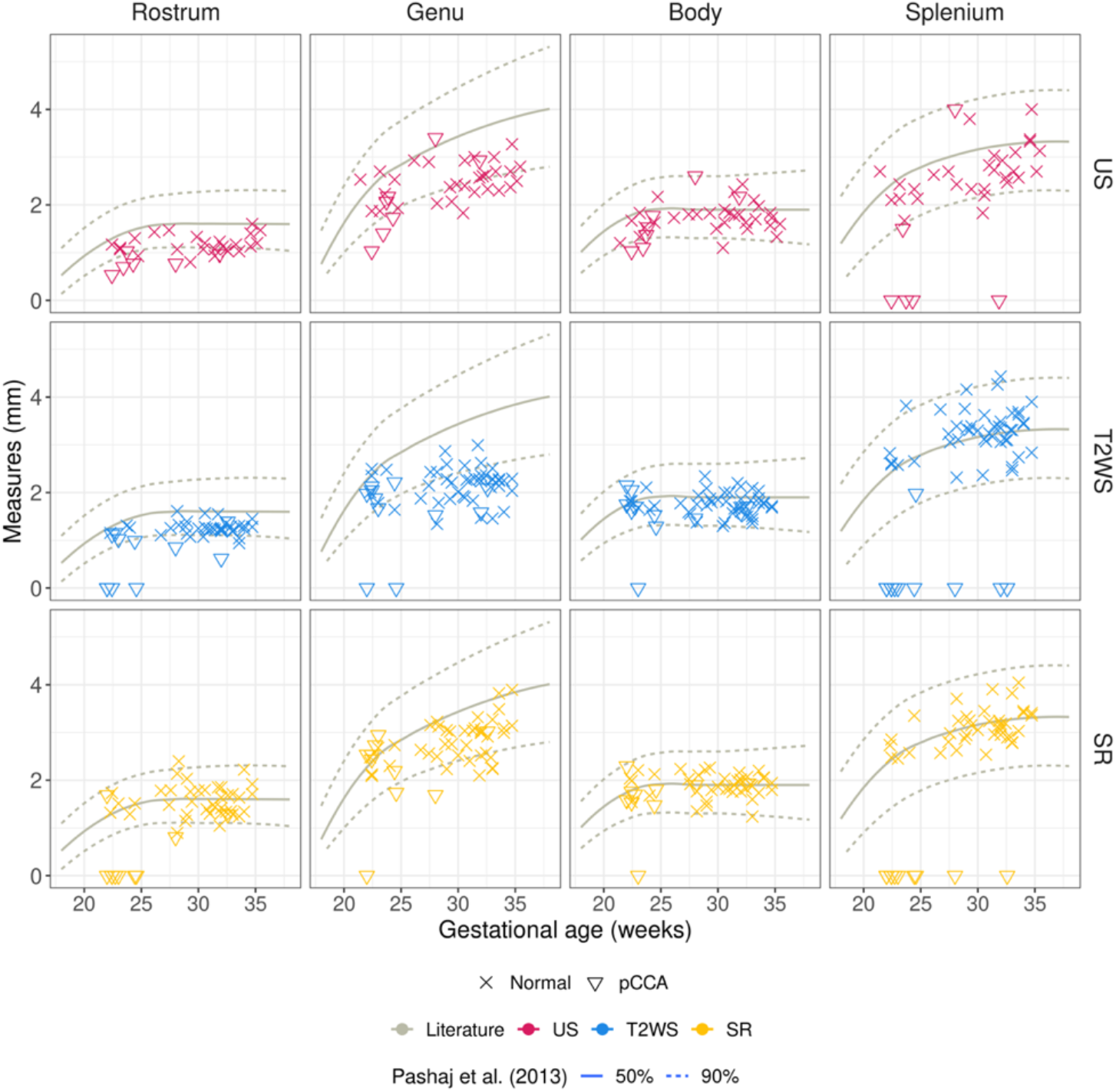
Analysis of CC heights in our cohort for US, T2WS and SR by obs1. Previous reported values in the literature (solid and dashed lines) are from US measurements reported in (31). Pathological subjects with partial CCA are shown (illustrated by triangles) but not used for the regression curves.

Intra-rater (obs1) statistical analysis of ***LCC*** and CC sub-segments’ heights measurements are reported in Table 2. Biometry differences between T2WS and SR were statistically significant for ***LCC***, ***Genu*** and ***Rostrum***. Between T2WS and US, statistically significant differences were found for the ***Genu*** and ***Splenium***. Finally, SR and US differences appear in ***LCC*** and ***Genu***.

**Table 1.** Percentage of measurements done per CC biometry

| % | LCC | Rostrum | Genu | Body | Splenium |
| --- | --- | --- | --- | --- | --- |
| **US** | 100 | 72 | 86 | 86 | 81 |
| **MR T2WS** | 100 | 82 | 96 | 100 | 98 |
| **MR SR** | 100 | 96 | 100 | 100 | 100 |

**Table 2.** Intra-observer (obs1) variability between the different imaging measurements of the CC length and the heights of its sub-segments.

| p-value<br>(sample size) | LCC | Rostrum | Genu | Body | Splenium |
| --- | --- | --- | --- | --- | --- |
| **US vs. T2WS** | 1.0<br>(N=43) | 1.0<br>(N=24) | **5.5e-03\***<br>(N=35) | 1.0<br>(N=37) | **6.7e-03 \***<br>(N=34) |
| **T2WS vs. SR** | 0.25<br>(N=51) | **2.7e-02\***<br>(N=42) | **1.1e-07\***<br>(N=49) | **2.7e-02\***<br>(N=51) | 0.41<br>(N=50) |
| **US vs. SR** | **1.4e-02\***<br>(N=38) | 0.32<br>(N=26) | **1.7e-02\***<br>(N=32) | 0.09<br>(N=32) | 0.14<br>(N=30) |

### 3.4 Expert US versus expert MRI

We first analyzed inter-modality performance minimizing the effect of the experience. Figure 7 shows the Bland-Altman plots of MR measures (expert is obs2) as compared to expert US (expert is obs1): negative differences mean MR measures overestimated US ones; average/median differences (solid lines) are around 0, but Rostrum is close to -1*mm*; few outliers (beyond dashed lines) are present. Additionally, regression of CC measurements with GA as compared with literature are shown in Supplementary Material (Figures S2 and S3). We explored whether there were statistically significantly different measurements between expert observers (Table 3). Results indicate that ***Rostrum*** has significantly different measurements between US and MR, both for T2WS and SR imaging, and ***LCC*** for US and SR only. Finally, we assessed the inter-observer (obs1 and obs2, junior and expert respectively) biometric measurements within MR SR imaging. They showed statistical differences (p < 0.05) for the heights of the ***Genu*** and the ***Rostrum*** (Table 4).

**Figure 7.**
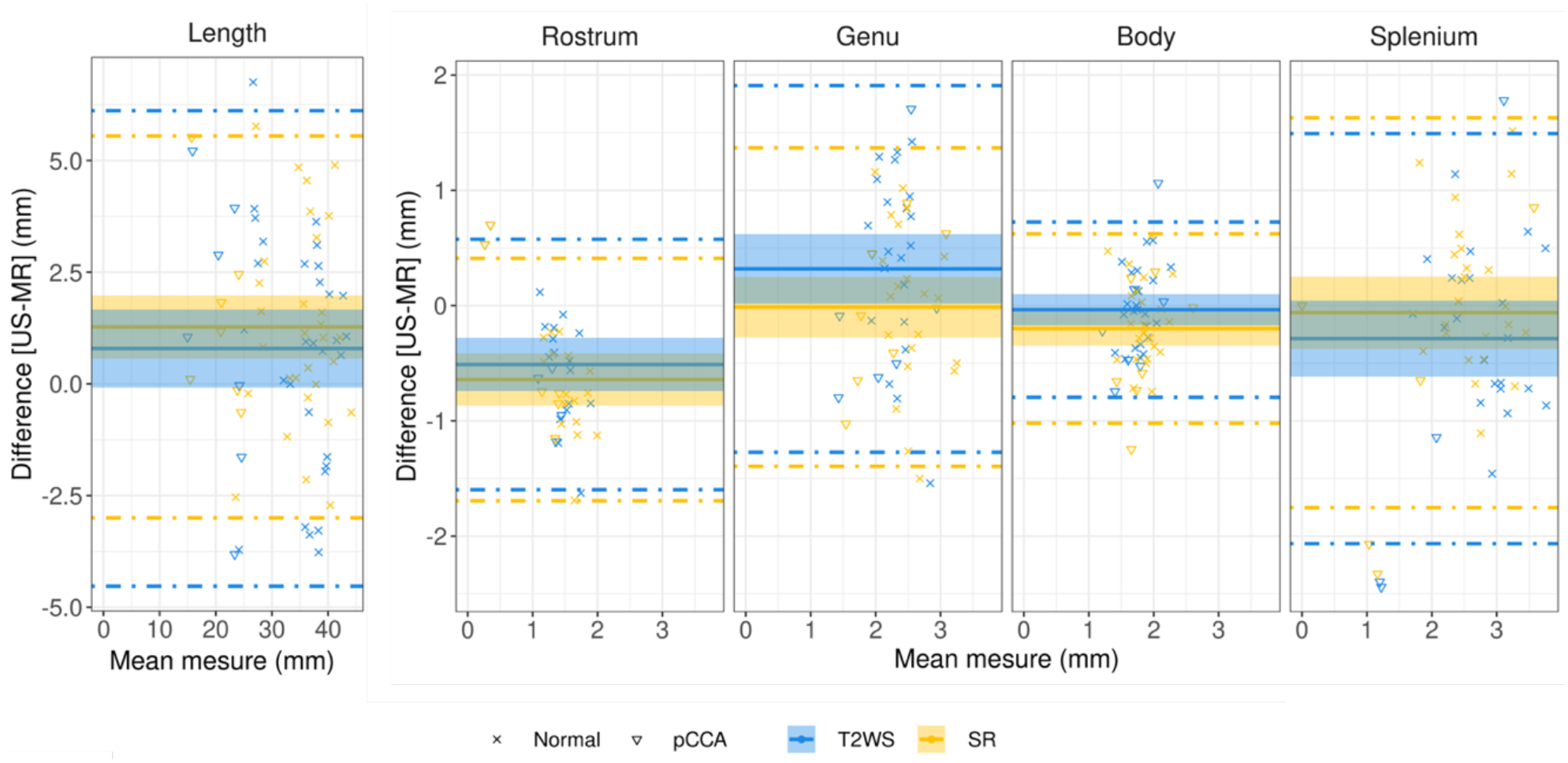
Bland-Altman plots for LCC and CC heights assessing expert US *vs* expert MRI measurements (T2WS in blue, SR in yellow) for all normal (cross symbol) and pathological subjects (triangle symbol) with partial CCA. The plot’s x-axis shows the average measurement, while the y-axis shows the difference in measurements between them. The average difference in measurements is represented by the solid line, while the 95 percent confidence interval limits are represented by the dashed lines. Dashed lines = 95% limits of agreement, and shadow areas correspond to the 95% confidence interval (CI).

**Table 3.** Statistical analysis inter-modality by experts US (Obs1) and MR (Obs2).

| p-value<br>(Sample size) | LCC | Rostrum | Genu | Body | Splenium |
| --- | --- | --- | --- | --- | --- |
| **US vs. T2WS** | 0.42<br>(N=42) | **1.6e-04\***<br>(N=28) | 7.7e-02<br>(N=34) | 1.0<br>(N=35) | 0.19<br>(N=34) |
| US vs. SR | 8.2e-03*<br>(N=38) | 9.1e-05*<br>(N=25) | 1.0<br>(N=31) | 6.6e-02<br>(N=32) | 1.0<br>(N=30) |

**Table 4.** Statistical analysis inter-observers within SR measurements.

| p-value<br>(Sample size) | LCC | Rostrum | Genu | Body | Splenium |
| --- | --- | --- | --- | --- | --- |
| SR | 1.0<br>(N=51) | 9e-05*<br>(N=48) | 1.1e-02*<br>(N=50) | 8.3e-02<br>(N=51) | 8.6e-02<br>(N=51) |

### 3.5 Area of the corpus callosum

Figure 8 shows our results of estimated CC area per GA. Previously reported results in the literature (33) from US are also illustrated (depicted in gray). Our results on US are in line with literature for young fetuses (<27 weeks of GA) but deviate more at later gestational stages (red). Interestingly, SR (yellow) mimics the same trend through GA than US, but slightly overestimates the area (same slope through GA). This might be due to higher partial volume effects and coarse spatial resolution in MR as compared to US. Different trend is depicted by area on T2WS (blue).

**Figure 8.**
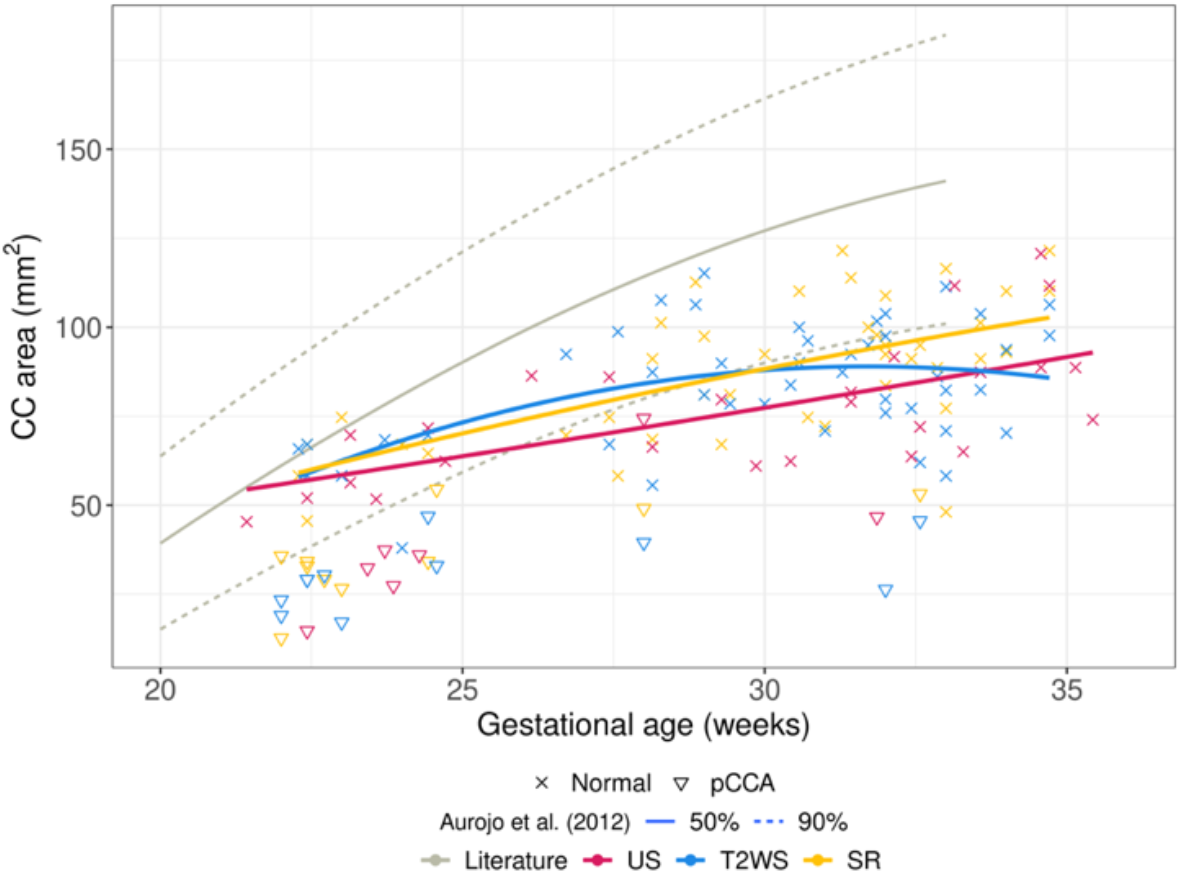
Area of CC: manual delineation on US, T2WS and SR. Pathological subjects with partial CCA are shown (illustrated by triangles) but not used for the regression curves.

## 4 DISCUSSION

### 4.1 The length of the CC can be accurately measured by both MR imaging methods

Several biometric charts of the fetal brain growth by US, including linear regressions of ***LCC***, have been published (29,31,34,35). Also, morphometry of the fetal brain by T2WS MRI has been previously explored (32,36,37). Besides, assessment of ***LCC*** measurements on SR in comparison to T2WS was presented in a recent study from our institution (22). Our first contribution is the study of ***LCC*** across gestational age (GA) on T2WS and SR on a larger cohort (around 50 subjects) and in comparison with expert US measurements, which consolidates the reliability of the previous works cited just above. Indeed, the ***LCC*** regression from SR measurements precisely overlays the US ones (both from our own cohort and from (31)) and slightly better than T2WS does, testifying of its proximity with the gold standard. Statistical paired-wise analysis of ***LCC*** measurements between US and SR (intra-observer 1 and inter-experts) is though significantly different. Givent though the excellent fit to US in the regression with GA, we hypothesize that this is due to the acquisition time difference between US and MRI, as evoked by the steep slope of the ***LCC*** growth.

### 4.2 Novel normative measurements of different CC segments heights across GA on MRI

Until today, fetal CC height measurements were only performed by mean of US imaging, as MRI spatial resolution was considered too low to provide trustable measures of such small structures (38,39). As such, no normative values for the different CC heights existed for fetal brain MRI, neither on clinical low-resolution series nor on super-resolution reconstructions. In our study, for the first time, we elaborated linear regressions and analyzed the reliability of MRI (T2WS and SR) in the measurement of the heights of the CC sub-segments. The CC heights on SR (and to a less extent T2WS) match well the US ones. All CC heights in both US and MR imaging settings fit well with the US ones reported previously in the literature (31), though to a lower extent the ***Genu***. Both observers in our study reported that SR helped for the assessment of the splenium, which is the most posterior part of the CC. We hypothesize that SR images of the fetal brain improve the visualization of the CC by dint of the enhanced through-plane spatial resolution that it offers and thanks to the opportunity to freely navigate in the 3 orthogonal views. This multiplanar approach diminishes the risk of confusion with other nearby anatomical structures of the CC whose MR image intensities are very close, such as the periventricular germ layer, the pericallosal and cingular sulci and the Galen’s and internal cerebral veins. This risk of error is in fact more pronounced if the images are oblique, a situation that may often occur when evaluating T2WS.

Currently, it is widely accepted that the quality of the MR imaging of the CC decreases in a postero-anterior fashion. Indeed, the more liquid the environment surrounding the CC is, as it is found in the posterior CC, the more contrast is created within the brain tissues, which ultimately allows for a better discrimination of the CC structure. As MR imaging mainly depends on the water content, while US imaging mainly depends on the impedance of the structures, this might explain why US is considered superior to MRI in the imaging of the anterior corpus callosum. Considering this, we postulated that SR MRI could help better visualize the whole CC. Our results show that the genu remains more difficult to see on MRI (both on T2WS and SR) compared to US. The lack of superiority of SR may be partially explained by the variable size of the broad genu according to the location of measurement, even if performed at the right designated anatomical location. Furthermore, as demonstrated by Pashaj et al (31), it is the segment whose biometry depends the most on the gestational age and, as such, can be the more sensitive CC sub-segment to the timeframe between the US et MRI acquisitions.

### 4.3 Less missing CC sub-segments measurements in SR MRI

In some cases, fetal brain MRI biometry may be useful in addition to the measures done in US, such as after an incomplete US examination due to an inconvenient maternal condition or an unfavorable position of the fetus (40). In our study, the rate of non-visualization of CC segments was lower on SR than on US and T2WS. Indeed, in SR, missing values were observed only in 2% of the exams versus 28.6% in US and 18.2% in T2WS. Most missing measurements were due to bad quality exams and concerned the rostrum. Although US is, in terms of heights measurements, the actual method of choice, it is user- and experience-dependent and highly reliant on the structure of interest. SR overcomes those issues by providing a unique 3D volume. The lower rate of missing values on SR compared with T2WS may also be explained by a greater confidence attributed to SR in comparison to T2WS in the identification of the CC, as it was already suggested in our previous study (22).

### 4.4 Inter-observer variability

The inter-experts’ analysis shows that, apart from ***LCC*** (for the reasons evoked in chapter 4.1), ***Rostrum*** is the only point of discrepancy between expert US and expert SR. This outcome is consistent with a previous observation from Garel *et al* (39), where the rostrum is considered as being very difficult to uncover, partially because of a thinner interhemispheric fissure and the absence of surrounding cerebro-spinal fluid and mainly because it is the thinnest part of the CC, measuring between 1 and 2 mm (41), while the resolution of SR is approximatively 1 mm^3^. This similarity between rostrum size and effective SR spatial resolution unveils the problem of the partial volume effect (12), which arises when more than one tissue type is present within a voxel, thus the different proportion of each tissue within that voxel contribute to the voxel intensity. Nevertheless, variability within SR measurements between observers reveals that both genu and rostrum heights are source of disagreement.

Overall, this study shows evidence that, although it is not made routinely, measuring the length, the middle and posterior CC sub-segments’ heights by the mean of SR MRI can have an interest in the clinical setting. Even if it needs further validation, it seems possible that in specific situations, such as when for some reason no good US is available, but a good MRI is, we could use it to measure the heights that are relevant for a handful of conditions such as such as CC hypo/hyperplasias or dysgenesis (38).

### 4.5 The role of the CC area

Few studies focused so far on the CC area and the reported surface of the CC and its sub-segments (33,35,42). Our CC area through gestation replicate only in part (for younger fetuses) previous reported results from Araujo et al (33). Interestingly, regression line of CC area from SR parallels the US curve, but overestimates the area, while the T2WS curve shows a parabolic trend. Smaller CC areas in abnormal development have been reported (43,44), paving the way of this biomarker for neurological outcome. With this diagnostical potential, reliable measurements of the area of the CC, such as provided by SR will become more and more important in the future. This will be especially relevant in the context of CC dysplasia, such as the thick/thin corpus callosum, that remain a difficult CC abnormality to diagnose with conventional methods.

### 4.6 Feasibility on patients with partial CC agenesis

We aimed also at exploring the feasibility of measurement on patients with partial CC agenesis (11 out of 57 fetuses in our cohort). Because of its bidirectional embryogenesis (45), when a default occurs during its period of formation and cause a pCCA, the most frequently missing sub-segments will be the most anterior and/or most posterior, respectively the rostrum or the splenium, which is well reflected in our pathological cohort. When measuring the CC sub-segments in a case of suspected pCCA, it is important to precisely respect the anatomical frontiers between the sub-segments, to identify whether a specific part of the CC is missing, as a whole or as a part. If they are integrated in our study, they though represent a small simple size that jeopardizes any sub-analysis specifically targeted on them. However, measurements on pCCA are still plotted on the regressions, and as expected they appear as clear outliers for ***LCC*** and with smaller measurements in the sub- segments (0 in case of absence). Future works need to target this population more specifically. Connecting prenatal biometry with post-natal clinical information in term of neurological development could definitely help clinician to perform a better pre-natal and post-natal counseling and management (20).

### 4.7 Limitations

Our study suffers from a relatively small cohort of subjects, nevertheless our results are in line with large scale US ((31) with sample size of 466) and MRI T2WS ((32) with sample size of 589 fetuses) studies. Additional limitations are the retrospective nature of our work and the lack of clinical and genetic follow-up (including neurological development in the early years of life and post-natal MRI). These limitations are shared by many studies on fetal brain biometry on US (35) as well as on MRI (36). However, following recent guidelines recommended in (35) for increasing methodology quality of fetal brain biometry studies on US, we elaborated numerical charts for the linear regressions, we were blinded for the gestational age, we report the inter- and intra-observer agreement and the non-visualization rate of the measurements. Another drawback is the two weeks’ timeframe set between the US et MRI imaging. However, it is highly accepted in the literature (MERIDIAN cohort (25)) and in our study the time difference between US and MR imaging acquisitions remains modest, with an average time difference between acquisitions of only of 3.9 days (range of 0 to 12 days). Moreover, the ratio of missing values in US images must be interpreted with caution as patients with a screening brain ultrasound that was not pathological do not necessarily have targeted CC images. Finally, gender effect was not evaluated but sexual dimorphism on fetal CC is still unclear, with studies that did (24) and did not show impact of gender (46).

### 4.8 Future directions of the research

Our work focuses on the assessment of SR MR imaging for CC measurements. Further work is still needed towards full automatization of existing SR pipelines and on reducing its computing time as to allow its integration into the clinical environment. The promise of a massive contribution of SR to diagnostic and therapy (and to the comprehension of the human neurological development) is though motivating (47).

We identified current spatial resolution and partial volume effect in MRI still limiting anterior CC measurements. A solution could be to thicken millimetric SR images, which may improve CC visibility in comparison with 1 mm^3^ SR, as it is routinely done daily on CT and MRI millimetric images, to search for brain metastasis for example, or on 3D fetal brain sonography for small areas, by manipulating the volume imaging contrast. This thicken strategy significantly increases the signal to noise ratio and image quality (48). Alternatively, SR reconstruction at finer spatial resolution scales could also be explored as to reduce the partial volume effects. Further, variability of measurements on different SR reconstruction pipelines could also be explored.

Today, the International Society of Ultrasound in Obstetrics and Gynecology (ISUOG) encourages 3T fetal MRI acquisitions which could provide higher resolution and signal to noise ratio (49). In our institution though, only around 30% of fetal examinations are done at 3T. Nevertheless, future research should evaluate the value of SR on CC assessment on 3T images and extend it to larger multi-centric cohort on normal and pCCA cases.

The addition of other imaging modalities and biomarkers would certainly support CC assessment. For instance diffusion MRI (50), MR spectroscopy (43) together with gyrification, neuropathological data (38), and long- term neurodevelopmental follow-up, might improve our understanding of the theories of formation of the CC and its neurological prognosis. This would allow a better classification of corpus callosum anomalies with an impact on prenatal counseling and management of patients. Moreover, the visualization of the other forebrain commissures may help refine the diagnosis better and define the commissural defects of the CC, such as the pCCA, distinguishing its various forms and of the other forebrain commissures (anterior commissure, hippocampal commissure).

## 5 CONCLUSIONS

This work shows that SR does not distort fetal CC biometry, providing with measurements close to US, except for its anterior part. Unlike US and T2WS, SR almost always allowed biometric measurements. In this context, we can encourage the performance of not only length, but also middle and posterior heights and surface area measurements by the mean of SR, at least when US is sub-optimal in the cases where CC anomalies are suspected. SR could be a turning point, combined with other advanced neuroimaging techniques, for a new classification of CC disruptions that could, alongside with genetic and tractography advances allow a better evaluation of the neurological prognosis, counseling, and therapy.

## DATA AVAILABILITY STATEMENT

The data analyzed in this study is subject to the following licenses/restrictions: the ethical approval for the use of these data did not include public release. Requests to access these datasets should be directed to Meritxell Bach Cuadra

## ETHICS STATEMENT

The studies involving human participants were reviewed and approved by the Commission cantonale (VD) d’éthique de la recherche sur l’être humain (CER-VD 2021-00124).

## Author Contributions

SL, IV, MK, MBC: conceptualization and design of the study; IV, LP, VD, MK: acquisition of data; SL, LP, IV, PGdD, VD, MK, MBC: analysis and interpretation of data; SL, LP, IV, MK: MRI/US measurements; IV, LP, MK, MBC: supervision; SL, PGdD, MK, MBC: writing—original draft; SL, PGdD, LP, IV, VD, MK, MBC: writing, revision, and editing of the submitted article.

## Data Availability

The data analyzed in this study is subject to the following licenses/restrictions: the ethical approval for the use of these data did not include public release. Requests to access these datasets should be directed to the authors.

## Acknowledgments

We acknowledge access to the facilities and expertise of the CIBM Center for Biomedical Imaging, a Swiss research center of excellence founded and supported by Lausanne University Hospital (CHUV), University of Lausanne (UNIL), Ecole Polytechnique Fédérale de Lausanne (EPFL), University of Geneva (UNIGE) and Geneva University Hospitals (HUG).

## REFERENCES

1. Leombroni M, Khalil A, Liberati M, D’Antonio F. Fetal midline anomalies: Diagnosis and counselling Part 1: Corpus callosum anomalies. European Journal of Paediatric Neurology. 2018 Nov;22(6):951–62.

2. Timor-Tritsch IE, Monteagudo A, Pilu G, editors. Ultrasonography of the prenatal brain. 3rd ed. New York: McGraw-Hill Professional; 2012. 490 p.

3. Griffiths PD, Brackley K, Bradburn M, Connolly DJA, Gawne-Cain ML, Griffiths DI, et al. Anatomical subgroup analysis of the MERIDIAN cohort: failed commissuration. Ultrasound Obstet Gynecol. 2017 Dec;50(6):753–60.

4. Raile V, Herz NA, Promnitz G, Schneider J, Tietze A, Kaindl AM. Clinical Outcome of Children With Corpus Callosum Agenesis. Pediatric Neurology. 2020 Nov;112:47–52.

5. Maurice P, Garel J, Garel C, Dhombres F, Friszer S, Guilbaud L, et al. New insights in cerebral findings associated with fetal myelomeningocele: a retrospective cohort study in a single tertiary centre. BJOG: Int J Obstet Gy. 2021 Jan;128(2):376–83.

6. The ENSO Working Group, Sileo FG, Pilu G, Prayer D, Rizzo G, Khalil A, et al. Role of prenatal magnetic resonance imaging in fetuses with isolated anomalies of corpus callosum: multinational study. Ultrasound in Obstet & Gyne. 2021 Jul;58(1):26–33.

7. Mahallati H, Sotiriadis A, Celestin C, Millischer AE, Sonigo P, Grevent D, et al. Heterogeneity in defining fetal corpus callosal pathology: systematic review. Ultrasound in Obstet & Gyne. 2021 Jul;58(1):11–8.

8. Malinger G, Paladini D, Haratz KK, Monteagudo A, Pilu GL, Timor-Tritsch IE. ISUOG Practice Guidelines (updated): sonographic examination of the fetal central nervous system. Part 1: performance of screening examination and indications for targeted neurosonography. Ultrasound Obstet Gynecol. 2020 Sep;56(3):476–84.

9. Paladini D, Malinger G, Pilu G, Timor-Trisch I, Volpe P. The MERIDIAN trial: caution is needed. The Lancet. 2017 May;389(10084):2103.

10. Bernardes da Cunha S, Carneiro MC, Miguel Sa M, Rodrigues A, Pina C. Neurodevelopmental Outcomes following Prenatal Diagnosis of Isolated Corpus Callosum Agenesis: A Systematic Review. Fetal Diagn Ther. 2021;48(2):88–95.

11. Pashaj S, Merz E. Detection of Fetal Corpus Callosum Abnormalities by Means of 3D Ultrasound. Ultraschall in Med. 2015 Nov 3;37(02):185–94.

12. Gholipour A, Rollins CK, Velasco-Annis C, Ouaalam A, Akhondi-Asl A, Afacan O, et al. A normative spatiotemporal MRI atlas of the fetal brain for automatic segmentation and analysis of early brain growth. Sci Rep. 2017 Mar 28;7(1):476.

13. Uus AU, Egloff Collado A, Roberts TA, Hajnal JV, Rutherford MA, Deprez M. Retrospective motion correction in foetal MRI for clinical applications: existing methods, applications and integration into clinical practice. BJR. 2022 Aug 8;20220071.

14. Gholipour A, Estroff JA, Warfield SK. Robust Super-Resolution Volume Reconstruction From Slice Acquisitions: Application to Fetal Brain MRI. Medical Imaging, IEEE Transactions on. 2010;29(10):1739–58.

15. Rousseau F, Glenn OA, Iordanova B, Rodriguez-Carranza C, Vigneron DB, Barkovich JA, et al. Registration-Based Approach for Reconstruction of High-Resolution In Utero Fetal MR Brain Images. Academic Radiology. 2006;13(9):1072–81.

16. Kuklisova-Murgasova M, Quaghebeur G, Rutherford MA, Hajnal JV, Schnabel JA. Reconstruction of fetal brain MRI with intensity matching and complete outlier removal. Medical Image Analysis. 2012;16(8):1550–64.

17. Tourbier S, Bresson X, Hagmann P, Thiran JP, Meuli R, Cuadra MB. An efficient total variation algorithm for super-resolution in fetal brain MRI with adaptive regularization. NeuroImage. 2015 Sep;118:584–97.

18. Kainz B, Steinberger M, Wein W, Kuklisova-Murgasova M, Malamateniou C, Keraudren K, et al. Fast volume reconstruction from motion corrupted stacks of 2D slices. Medical Imaging, IEEE Transactions on. 2015 Sep;34(9):1901–13.

19. Ebner M, Wang G, Li W, Aertsen M, Patel PA, Aughwane R, et al. An automated framework for localization, segmentation and super-resolution reconstruction of fetal brain MRI. NeuroImage. 2020 Feb;206:116324.

20. Pier DB, Gholipour A, Afacan O, Velasco-Annis C, Clancy S, Kapur K, et al. 3D Super-Resolution Motion-Corrected MRI: Validation of Fetal Posterior Fossa Measurements: Super-Resolution Motion- Corrected MRI of the Fetal Posterior Fossa. Journal of Neuroimaging. 2016 Sep;26(5):539–44.

21. Kyriakopoulou V, Vatansever D, Davidson A, Patkee P, Elkommos S, Chew A, et al. Normative biometry of the fetal brain using magnetic resonance imaging. Brain Structure and Function. 2017 Jul;222(5):2295–307.

22. Khawam M, De Dumast P, Deman P, Kebiri H, Yu T, Tourbier S, et al. Fetal brain biometric measurements on 3D super-resolution reconstructed T2-weighted MRI: An intra-and inter-observer agreement study. Frontiers in pediatrics. 2021;9:639746.

23. Velasco-Annis C, Gholipour A, Afacan O, Prabhu SP, Estroff JA, Warfield SK. Normative biometrics for fetal ocular growth using volumetric MRI reconstruction: Normative biometrics for fetal ocular growth. Prenatal Diagnosis. 2015 Apr;35(4):400–8.

24. Machado-Rivas F, Gandhi J, Choi JJ, Velasco-Annis C, Afacan O, Warfield SK, et al. Normal Growth, Sexual Dimorphism, and Lateral Asymmetries at Fetal Brain MRI. Radiology. 2022 Apr;303(1):162–70.

25. Griffiths PD, Bradburn M, Campbell MJ, Cooper CL, Graham R, Jarvis D, et al. Use of MRI in the diagnosis of fetal brain abnormalities in utero (MERIDIAN): a multicentre, prospective cohort study. The Lancet. 2017 Feb;389(10068):538–46.

26. Garcia-Flores J, Recio M, Uriel M, Cañamares M, Cruceyra M, Tamarit I, et al. Fetal magnetic resonance imaging and neurosonography in congenital neurological anomalies: supplementary diagnostic and postnatal prognostic value. The Journal of Maternal-Fetal & Neonatal Medicine. 2013 Oct;26(15):1517–23.

27. The ENSO Working Group, Di Mascio D, Khalil A, Thilaganathan B, Rizzo G, Buca D, et al. Role of prenatal magnetic resonance imaging in fetuses with isolated mild or moderate ventriculomegaly in the era of neurosonography: international multicenter study. Ultrasound Obstet Gynecol. 2020 Sep;56(3):340–7.

28. 1. Basics of 3D and 4D Volume Acquisition. In: 3D Ultrasound in Prenatal Diagnosis [Internet]. De Gruyter; 2016 [cited 2023 Mar 17]. p. 3–14. Available from: https://www.degruyter.com/document/doi/10.1515/9783110497359-002/html

29. Pomar L, Baert J, Mchirgui A, Lambert V, Carles G, Hcini N, et al. Comparison between Two-Dimensional and Three-Dimensional Assessments of the Fetal Corpus Callosum: Reproducibility of Measurements and Acquisition Time. Journal of Pediatric Neurology. 2021 Oct;19(05):312–20.

30. Yushkevich PA, Piven J, Hazlett HC, Smith RG, Ho S, Gee JC, et al. User-guided 3D active contour segmentation of anatomical structures: Significantly improved efficiency and reliability. NeuroImage. 2006 Jul;31(3):1116–28.

31. Pashaj S, Merz E, Wellek S. Biometry of the fetal corpus callosum by three-dimensional ultrasound: Biometry of the fetal corpus callosum. Ultrasound Obstet Gynecol. 2013 Dec;42(6):691–8.

32. Tilea B, Alberti C, Adamsbaum C, Armoogum P, Oury JF, Cabrol D, et al. Cerebral biometry in fetal magnetic resonance imaging: new reference data. Ultrasound in Obstetrics and Gynecology. 2009 Feb;33(2):173–81.

33. Júnior EA, Visentainer M, Simioni C, Ruano R, Nardozza LMM, Moron AF. Reference Values for the Length and Area of the Fetal Corpus Callosum on 3-Dimensional Sonography Using the Transfrontal View. Journal of Ultrasound in Medicine. 2012 Feb;31(2):205–12.

34. Achiron R, Achiron A. Development of the human fetal corpus callosum: a high-resolution, cross-sectional sonographic study: Fetal corpus callosum development. Ultrasound Obstet Gynecol. 2001 Oct;18(4):343–7.

35. Rosenbloom JI, Yaeger LH, Porat S. Reference Ranges for Corpus Callosum and Cavum Septi Pellucidi Biometry on Prenatal Ultrasound: Systematic Review and Meta-Analysis. J of Ultrasound Medicine. 2022 Sep;41(9):2135–48.

36. Di Mascio D, Khalil A, Rizzo G, Kasprian G, Caulo M, Manganaro L, et al. Reference ranges for fetal brain structures using magnetic resonance imaging: systematic review. Ultrasound in Obstet & Gyne. 2022 Mar;59(3):296–303.

37. Harreld JH, Bhore R, Chason DP, Twickler DM. Corpus Callosum Length by Gestational Age as Evaluated by Fetal MR Imaging. AJNR Am J Neuroradiol. 2011 Mar;32(3):490–4.

38. Izzo G, Toto V, Doneda C, Parazzini C, Lanna M, Bulfamante G, et al. Fetal thick corpus callosum: new insights from neuroimaging and neuropathology in two cases and literature review. Neuroradiology. 2021 Dec;63(12):2139–48.

39. Garel C, Cassart M. Imagerie du fœtus au nouveau-né. 2016.

40. Prayer D, Malinger G, Brugger PC, Cassady C, De Catte L, De Keersmaecker B, et al. ISUOG Practice Guidelines: performance of fetal magnetic resonance imaging. Ultrasound in Obstet & Gyne. 2017 May;49(5):671–80.

41. Raybaud C. The corpus callosum, the other great forebrain commissures, and the septum pellucidum: anatomy, development, and malformation. Neuroradiology. 2010 Jun;52(6):447–77.

42. Shi Y, Xue Y, Chen C, Lin K, Zhou Z. Association of gestational age with MRI-based biometrics of brain development in fetuses. BMC Med Imaging. 2020 Dec;20(1):125.

43. Sanz-Cortes M, Egaña-Ugrinovic G, Simoes RV, Vazquez L, Bargallo N, Gratacos E. Association of brain metabolism with sulcation and corpus callosum development assessed by MRI in late-onset small fetuses. American Journal of Obstetrics and Gynecology. 2015 Jun;212(6):804.e1–804.e8.

44. Egaña-Ugrinovic G, Sanz-Cortés M, Couve-Pérez C, Figueras F, Gratacós E. Corpus callosum differences assessed by fetal MRI in late-onset intrauterine growth restriction and its association with neurobehavior: Corpus callosum assessment in term IUGR and its correlation with neurobehavior. Prenat Diagn. 2014 Sep;34(9):843–9.

45. Birnbaum R, Barzilay R, Brusilov M, Wolman I, Malinger G. The early pattern of human corpus callosum development: A transvaginal 3D neurosonographic study. Prenatal Diagnosis. 2020 Sep;40(10):1239–45.

46. Gafner M, Kedar Sade E, Barzilay E, Katorza E. Sexual dimorphism of the fetal brain biometry: an MRI-based study. Arch Gynecol Obstet [Internet]. 2022 Oct 17 [cited 2023 Mar 18]; Available from: https://link.springer.com/10.1007/s00404-022-06818-4

47. Stout JN, Bedoya MA, Grant PE, Estroff JA. Fetal Neuroimaging Updates. Magnetic Resonance Imaging Clinics of North America. 2021 Nov;29(4):557–81.

48. Paladini D, Malinger G, Birnbaum R, Monteagudo A, Pilu G, Salomon LJ, et al. ISUOG Practice Guidelines (updated): sonographic examination of the fetal central nervous system. Part 2: performance of targeted neurosonography. Ultrasound in Obstet & Gyne. 2021 Apr;57(4):661–71.

49. Cassart M, Garel C. European overview of current practice of fetal imaging by pediatric radiologists: a new task force is launched. Pediatr Radiol. 2020 Nov;50(12):1794–8.

50. Millischer AE, Grevent D, Sonigo P, Bahi-Buisson N, Desguerre I, Mahallati H, et al. Feasibility and Added Value of Fetal DTI Tractography in the Evaluation of an Isolated Short Corpus Callosum: Preliminary Results. AJNR Am J Neuroradiol. 2022 Jan;43(1):132–8.

