## Supplementary material for "Assessment of fetal corpus callosum biometry by 3D super-resolution reconstructed T2-weighted MRI"

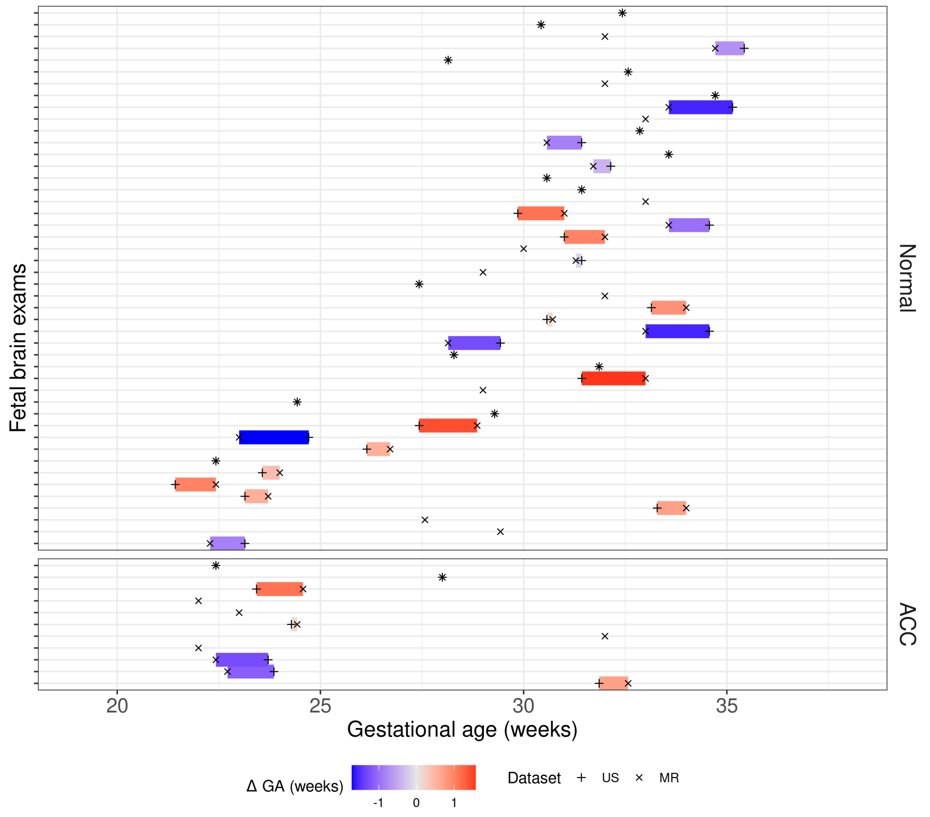


**Figure S1** Gestational age at moment of Ultrasound (+) and MR (x) acquisition. Dark colors (blue or red) illustrate the largest time difference.

**Table S1** Intra class correlation coefficient (ICC) for observer 1. Average on 9 normal and 3 pCCA repeated measurements.

| ICC (Obs1) | US | T2WS | SR |
|---|---|---|---|
| **LCC** | 0.999 | 0.996 | 0.995 |
| **Splenium** | 0.979 | 0.993 | 0.979 |
| **Body** | 0.832 | 0.707 | 0.145 |
| **Genu** | 0.821 | 0.812 | 0.536 |
| **Rostrum** | 0.887 | 0.903 | 0.579 |
| **Overall** | 1 | 0.99 | 0.99 |

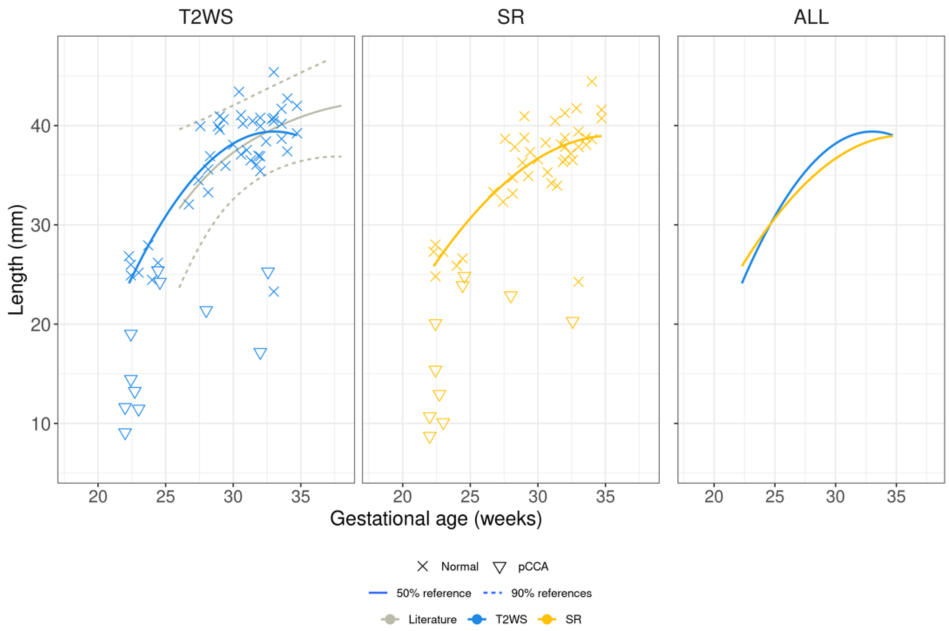


**Figure S2** Regression of **LCC** measurements of Obs2 (expert MR) with gestational age.

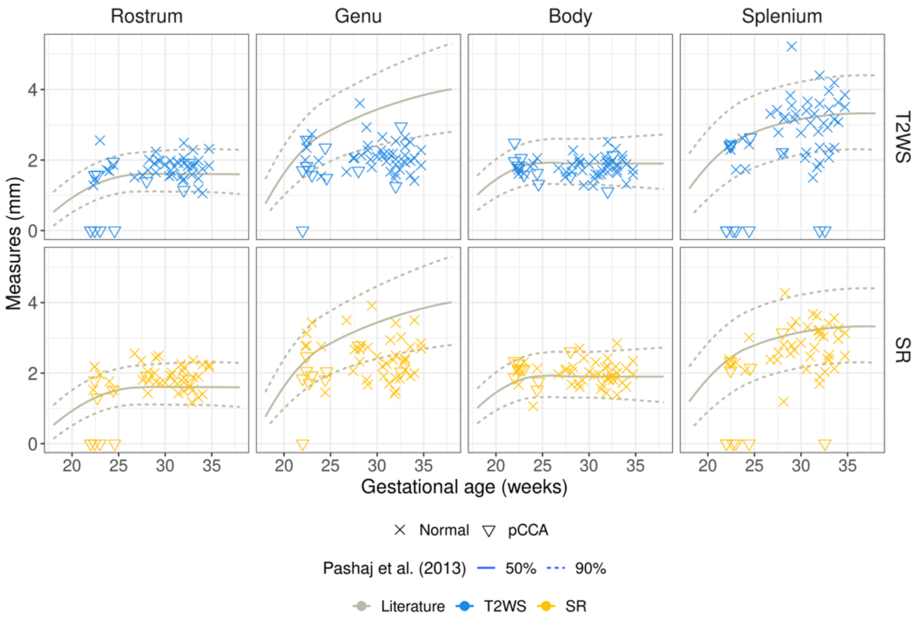


**Figure S3** Regression of CC heights measurements (Rostrum, Genu, Body and Splenium) of Obs2 (expert MR) with gestational age.
